# Prevalence, Morbidity, and Mortality of 1,609 Men with Sex Chromosome Aneuploidy: Results from the Diverse Million Veteran Program Cohort

**DOI:** 10.1101/2023.07.15.23292710

**Authors:** Shanlee M. Davis, Craig Teerlink, Julie A. Lynch, Bryan R. Gorman, Meghana Pagadala, Aoxing Liu, Matthew S. Panizzon, Victoria C. Merritt, Giulio Genovese, Saiju Pyarajan, Judith L. Ross, Richard L. Hauger

**Author notes:** **Corresponding Author:** Shanlee Davis, MD, PhD; 13123 East 16^th^ Ave B265, Aurora, CO 80045, Ph. Joint first authors (Shanlee M. Davis, Craig Teerlink). Joint senior authors (Judith L. Ross, Richard L. Hauger).

## Abstract

**Importance:** The reported phenotypes of men with 47,XXY and 47,XYY syndromes include tall stature, multisystem comorbidities, and poor health-related quality of life (HRQoL). However, knowledge about these sex chromosome aneuploidy (SCA) conditions has been derived from studies in the <15% of patients who are clinically diagnosed and also lack diversity in age and genetic ancestry.

**Objectives:** Determine the prevalence of clinically diagnosed and undiagnosed X or Y chromosome aneuploidy among men enrolled in the Million Veteran Program (MVP); describe military service metrics of men with SCAs; compare morbidity and mortality outcomes between men with SCA with and without a clinical diagnosis to matched controls.

**Design:** Cross-sectional, case-control

**Setting:** United States Veterans Administration Healthcare System

**Participants:** Biologic males enrolled in the MVP biobank with genomic identification of an additional X or Y chromosome (cases); controls matched 1:5 on sex, age, and genetic ancestry

**Main Outcome(s) and Measure(s):** Prevalence of men with SCAs from genomic analysis; clinical SCA diagnosis; Charlson Comorbidity Index (CCI); rates of outpatient, inpatient, and emergency encounters per year; self-reported health outcomes; standardized mortality ratio (SMR)

**Results:** An additional X or Y chromosome was present in 145 and 125 per 100,000 males in the MVP, respectively, with the highest prevalence among men with European and East Asian ancestry. At a mean age of 61±12 years, 74% of male veterans with 47,XXY and >99% with 47,XYY remained undiagnosed. Individuals with 47,XXY (n=862) and 47,XYY (n=747) had similar military service history, all-cause SMR, and age of death compared to matched controls. CCI and healthcare utilization were higher among individuals with SCA, while several measures of HRQoL were lower. Men with a clinical diagnosis of 47,XXY had higher healthcare utilization but lower comorbidity score compared to those undiagnosed.

**Conclusion and Relevance:** One in 370 males in the MVP cohort have SCA, a prevalence comparable to estimates in the general population. While these men have successfully served in the military, they have higher morbidity and report poorer HRQoL with aging. Longer longitudinal follow-up of this sample will be informative for clinical and patient-reported outcomes, the role of ancestry, and mortality statistics.

**KEY POINTS:**

1. Comparable to the general population, approximately 1 in 370 male veterans have a sex chromosome aneuploidy, but most are undiagnosed.
2. Men with X or Y chromosome aneuploidy successfully complete US miliary duty with similar service history compared to their 46,XY peers.
3. Medical comorbidities and healthcare utilization metrics are higher in male veterans with 47,XXY and 47,XYY during aging, however life expectancy is similar to matched controls.

## INTRODUCTION

Approximately 1 in 400 males have an extra X or Y chromosome based on birth cohort studies, resulting in ∼5,000 males born in the US annually with 47,XXY (Klinefelter syndrome) or 47,XYY (Double Y syndrome).^1^ These male sex chromosome aneuploidy (SCA) conditions are associated with broad multisystem medical and neuropsychiatric comorbidities and certain physical traits such as tall stature; however, there is substantial phenotypic variability and lack of pathognomonic dysmorphic features, resulting in profound underdiagnosis with <25% of men with 47,XXY and <10% with 47,XYY receiving a diagnosis.^1^ With few exceptions, SCA research has been limited to the minority of individuals who have a clinical diagnosis, leading to substantial ascertainment bias. Therefore, the full spectrum of health outcomes for individuals with 47,XXY or 47,XYY is unknown.

British and Danish population registries have informed most of our knowledge about health outcomes for individuals with SCA conditions. These studies report a decreased life span and higher incidence rates for nearly all clinical diagnostic codes in males clinically diagnosed with SCA compared to sex- and age-matched controls.^2-6^ In addition to increased morbidity and mortality outcomes, a recent meta-analysis summarizing the published literature on quality of life (QoL) in men with 47,XXY (13 studies with a total of 829 participants) found a negative impact on physical, psychological, and social QoL domains.^7^ However, these studies only reflect men with a clinical SCA diagnosis and therefore cannot be generalized to all individuals with SCA. Recently, a study using the UK Biobank identified 356 men via genotype array to have X or Y chromosome polysomy.^8^ While the prevalence of 47,XXY and 47,XYY was lower than previous estimates, most of their findings aligned with prior data, including an 85% non-diagnosis rate and poorer overall health outcomes compared to men with 46,XY from both self-report and medical record data sources. A significant limitation, however is that epidemiological data and most clinical trial data in SCA conditions have been based on men with European ancestry. While Western European countries have the advantage of national healthcare systems that make epidemiological research more feasible, they lack the unique ethnic and socioeconomic diversity present in the US and other parts of the world. In addition, there is a paucity of data exploring how SCA affects biological aging and the development of age-related medical disorders.

The Veteran’s Health Administration (VA) Million Veteran Program (MVP) is a voluntary population-based study of genetic determinants of medical, neurological, and psychiatric illnesses and health outcomes for individuals who have served in the US military.^9^ We used the MVP cohort as a novel opportunity to address limitations in existing 47,XXY and 47,XYY research. The aims of our investigation were to 1) determine the prevalence of men in MVP with X or Y chromosome aneuploidy overall, both with and without a clinical diagnosis, 2) describe military service for men with SCAs compared to matched controls, and 3) compare morbidity and mortality outcomes between men with SCAs to matched controls, as well as between those with and without a clinical diagnosis.

## METHODS

### Overview of study design

The MVP began enrollment in 2011 and will soon have actively recruited 1 million participants across multiple VA facilities within the US. Methodological details for MVP have been described at length elsewhere.^9^ Veterans enrolled in MVP consent to provide a blood sample used for genotyping and biobanking, grant access to their VA electronic health record (EHR) for research, and complete the Baseline Survey and Lifestyle Survey.^10^ The MVP study was approved by the VA Central Institutional Review Board (IRB), and all individuals provided informed consent prior to study enrollment. This cross-sectional, case-control study analyzed genotype, EHR, and survey responses of 658,582 men with both genotype and VA EHR data available (Figure 1a). Survey response rates in our cohort were similar to MVP metrics as a whole.^11^

**Figure 1.**
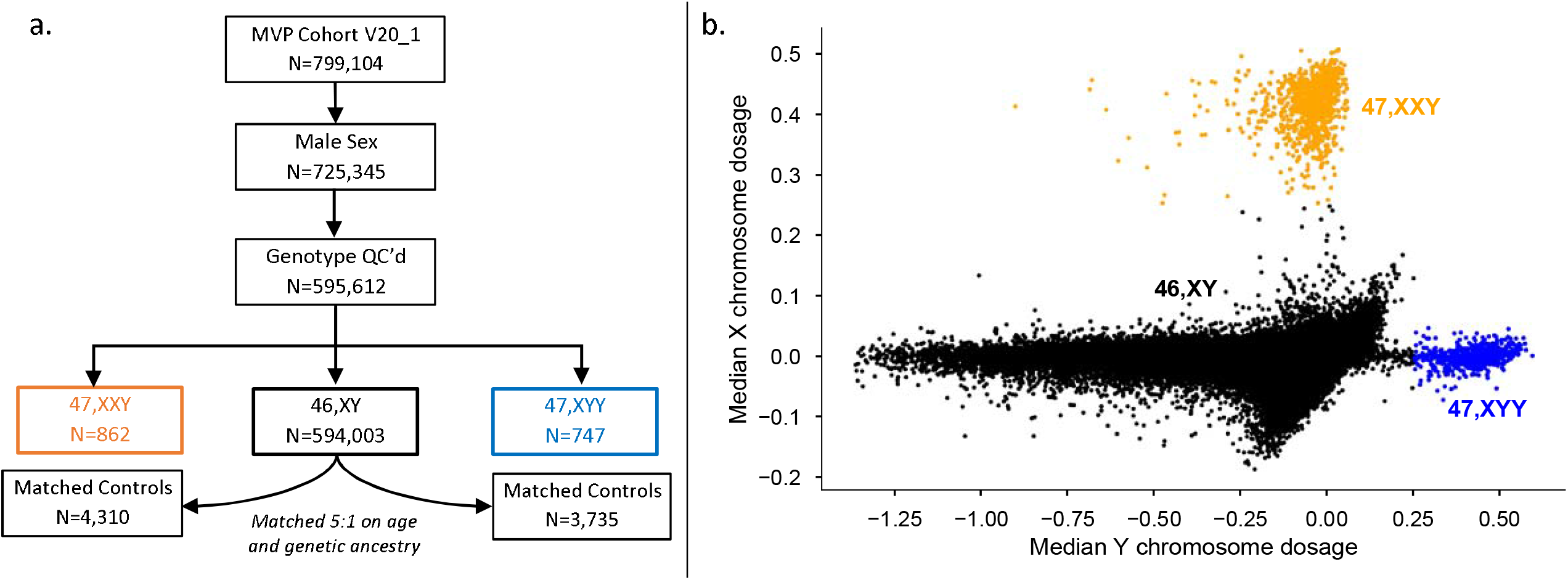
(a) Flow diagram of MVP participants included in the current analysis. (b) Scatterplot of sex chromosome array intensity over >20,000 X probes and 157 Y probes in all male MVP participants to identify men with an additional X or Y chromosome. Orange dots are males with 47,XXY, blue dots are males with 47,XYY, and black are males with 46,XY.

### Genotyping, Ancestry, and SCA determination

Blood samples are banked at the VA Central Biorepository at the Massachusetts Veterans Epidemiology Research and Information Center (MAVERIC). DNA is extracted from peripheral blood leukocytes and analyzed on the MVP custom Affymetrix Axiom Biobank 1.0 array consisting of 723,305 unique SNPs. Details on this platform, technical processes, and quality control measures have previously been published.^12^ Genotype data within the GenISIS workspace were used to determine genetic ancestry using the Harmonized Ancestry and Race/Ethnicity (HARE) method.^13^ To identify men with an additional X or Y chromosome, the sex chromosome dosage was estimated by array intensity measurements. Normalized probeset intensities (log-R ratios) were calculated over 20,170 non-pseudoautosomal X probesets and 157 non-pseudoautosomal Y probesets. Comparison of median log-R ratios on X and Y (mLRR-X and mLRR-Y) identified clear clusters corresponding to samples with 46,XX, 46,XY, 47,XXY and 47,XYY karyotypes. A final SCA call set was then obtained by thresholding mLRR-X and mLRR-Y values around each identified cluster (Figure 1b).

### Analytic Cohort

Individuals with available VA EHR and male sex who were within the 47,XXY or 47,XYY clusters defined our cases for this analysis, while those in the 46,XY cluster were controls. Analyses were not stratified by race because presence of an additional sex chromosome was considered as a single genetic variant eliminating the possibility of population stratification due to genetic drift influencing statistical tests, therefore we proceeded with a matched analysis where each male with an extra X or Y chromosome was randomly matched with five 46,XY males on age at time of enrollment and ancestry based on HARE designation.^14^

### Outcomes

We determined the prevalence of SCA based on the number of males with a second X or Y chromosome over the total number of males in MVP expressed as the number per 100,000 males, and then stratified by genetic ancestry. To determine whether an individual with a genetically identified 47,XXY or 47,XYY karyotype had a clinical diagnosis of SCA, we queried ICD diagnosis codes consistent with SCA (International Classification of Disease 9 758.7-81; ICD10 Q98.0-9). Participants with at least one diagnostic code were determined to be clinically diagnosed, and the age first diagnosis code was retrieved.

Descriptive military service metrics were obtained from a combination of VA records (e.g. branch, time period, discharge status) and survey responses (e.g. active duty, deployment, exposures). We calculated healthcare utilization metrics including the number of outpatient, emergency, and overnight hospital encounters per year followed, with duration of follow-up determined as the difference between the initial and most recent encounter dates. The primary measure of medical morbidity was the Charlson Comorbidity Index (CCI), a calculated metric incorporating multiple medical conditions obtained from the EHR validated to estimate 10-year survival.^15^ Additional measures of morbidity included body mass index (BMI) from EHR, self-reported number of prescription medications, and validated summary scores from the Baseline and Lifestyle surveys as previously described^10^ including the Veterans RAND 12 Item Health Survey (VR-12)^16,17^ as a measure of physical and mental health-related QoL, Medical Outcomes Study Cognitive Functioning-Revised Scale (MOS-Cog-R)^18,19^ as a subjective estimate of cognitive difficulties, and the Patient Health Questionnaire-4 (PHQ-4)^20^ and the Posttraumatic Stress Disorder Checklist (PCL)^21^ as estimates of psychological functioning.

Finally, we calculated the standardized mortality ratio (SMR) as the number of deaths in cases divided by the expected number of deaths based on matched controls within the same time period. The primary cause of death was recoded into one of 39 cause of death categories according to the National Center for Health Statistics manual.^22^

### Analytic approach

Continuous variables are reported as means and standard deviations or medians with interquartile range; categorical variables are reported as frequency/number and percentage. We conducted outcome comparisons between cases (47,XXY or 47,XYY) versus their matched controls using chi-squared tests for categorical variables and t-tests for continuous variables. Similarly, we used chi-squared and t-tests to compare those with and without a clinical SCA diagnosis. To avoid type 1 error with a large sample size and multiple comparisons, we elected a conservative alpha of 0.005 for primary outcomes. Analyses were conducted using R v4.0.3 and Prism Graphpad v9.5.1.

## RESULTS

Out of 595,612 genotyped males, we identified 862 with an additional X chromosome (47,XXY) and 747 with an extra Y chromosome (47,XYY), corresponding to a prevalence of 145 per 100,000 (1 in 690) males for 47,XXY and 125 per 100,000 (1 in 800) males for 47,XYY (Figure 1). The average age of enrollment was similar to the overall MVP male population; however, genetic ancestry was significantly different (Table 1; Figure 2). This resulted in an estimated prevalence over twice as high in East Asian and European ancestry groups compared to those of African or Hispanic (p<0.001). The minority of men with 47,XXY (n=226, 26%) had a diagnosis in their EHR consistent with a clinical diagnosis of Klinefelter syndrome, and only two men with 47,XYY had a diagnosis consistent with clinical ascertainment.

**Table 1.** Demographics and Prevalence of Males with Sex Chromosome Aneuploidy Within the MVP Population.

|  |  |  |  |
| --- | --- | --- | --- |
|  | **All males in**<br>**the MVP**<br>**N=595,612** | **Males with**<br>**47,XXY**<br>**N=862** | **Males with**<br>**47,XYY**<br>**N=747** |
| Age at enrollment (yrs) | 61.5 ± 14.2<br>64 (18-106) | 61.1 ± 12.0<br>63 (22-91) | 60.7 ± 12.2<br>63 (26-97) |
| Genetic Ancestry<sup>^</sup> |  |  |  |
| African | 105,842 (17.7%) | 87 (10.1%) | 46 (6.2%) |
| East Asian | 7,313 (1.2%) | 10 (1.2%) | 14 (1.9%) |
| European | 427,143 (71.8%) | 725 (84.1%) | 625 (83.7%) |
| Hispanic | 46,828 (7.8%) | 32 (3.7%) | 53 (7.1%) |
| Unavailable | 8,486 (1.4%) | 8 (0.9%) | 9 (1.2%) |
| Clinical SCA diagnosis | 228 (0.04%) | 226 (26.2%) | 2 (0.3%) |
| Age first documented | 52 (22-85) | 52 (22-85) | 59 (49, 68) |
| <sup>^</sup>Chi squared comparisons of ancestry categories for 47,XXY vs all MVP and 47,XYY vs all MVP are both p<0.0001<br>Data are shown as mean ± standard deviation (SD) and/or median (range) for numerical data, n (%) for categorical data. MVP = Million Veteran Program; SCA = sex chromosome aneuploidy |  |  |  |

**Figure 2.**
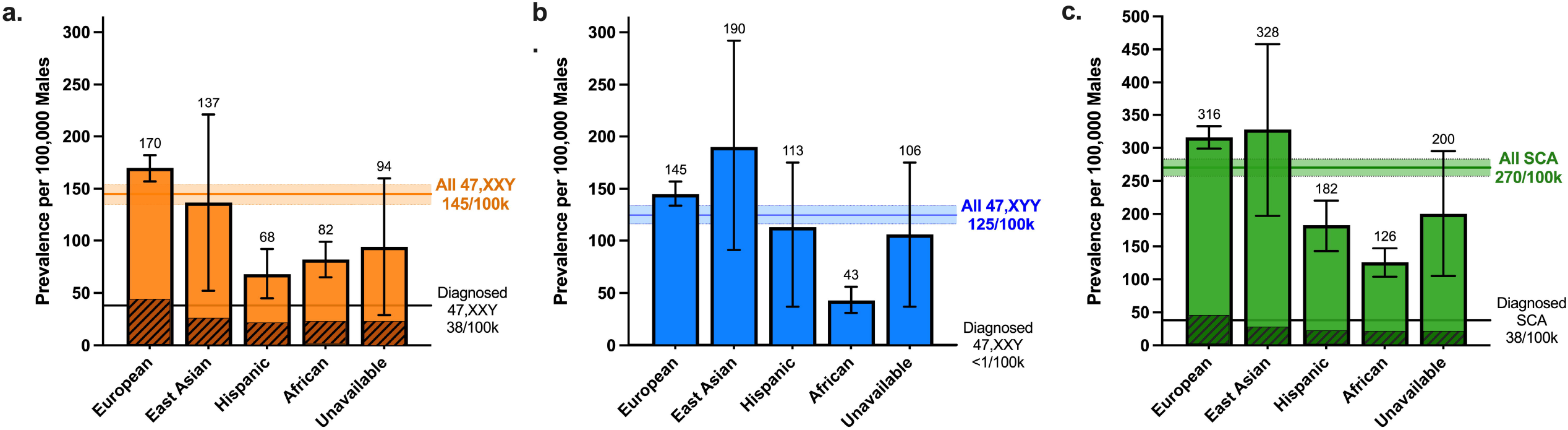
Prevalence of (a) 47,XXY, (b) 47,XYY, and (c) both 47,XXY and 47,XYY combined per 100,000 males in the MVP stratified by genetic ancestry. The bars and error bars represent the calculated prevalence and 95% confidence interval (CI) for each genetic ancestry group. The horizontal line and lighter shaded area represent the calculated prevalence and 95% CI overall. The darker shaded area within the bars represent those with a clinical diagnosis in their electronic medical record.

After matching on age and ancestry, military service metrics were very similar between SCA cases and controls and nearly all (>90%) earned an Honorable Discharge (Table 2). Men with 47,XXY and 47,XYY were 4.6 and 9.2cm taller than controls respectively (p<0.001). CCI-calculated 10-year survival estimate mean was 44% in 47,XXY and 39% in 47,XYY vs ∼57% in controls (p<0.001). The rates of outpatient, emergency, and inpatient encounters within the VA system were 25-50% higher for SCA groups compared to controls (p<0.001 for all). Similarly, men with SCAs self-reported significantly more hospitalizations, prescription medications, and reproductive and neurological health concerns than matched controls on survey responses, along with lower health-related QoL (Table 3).

**Table 2.**
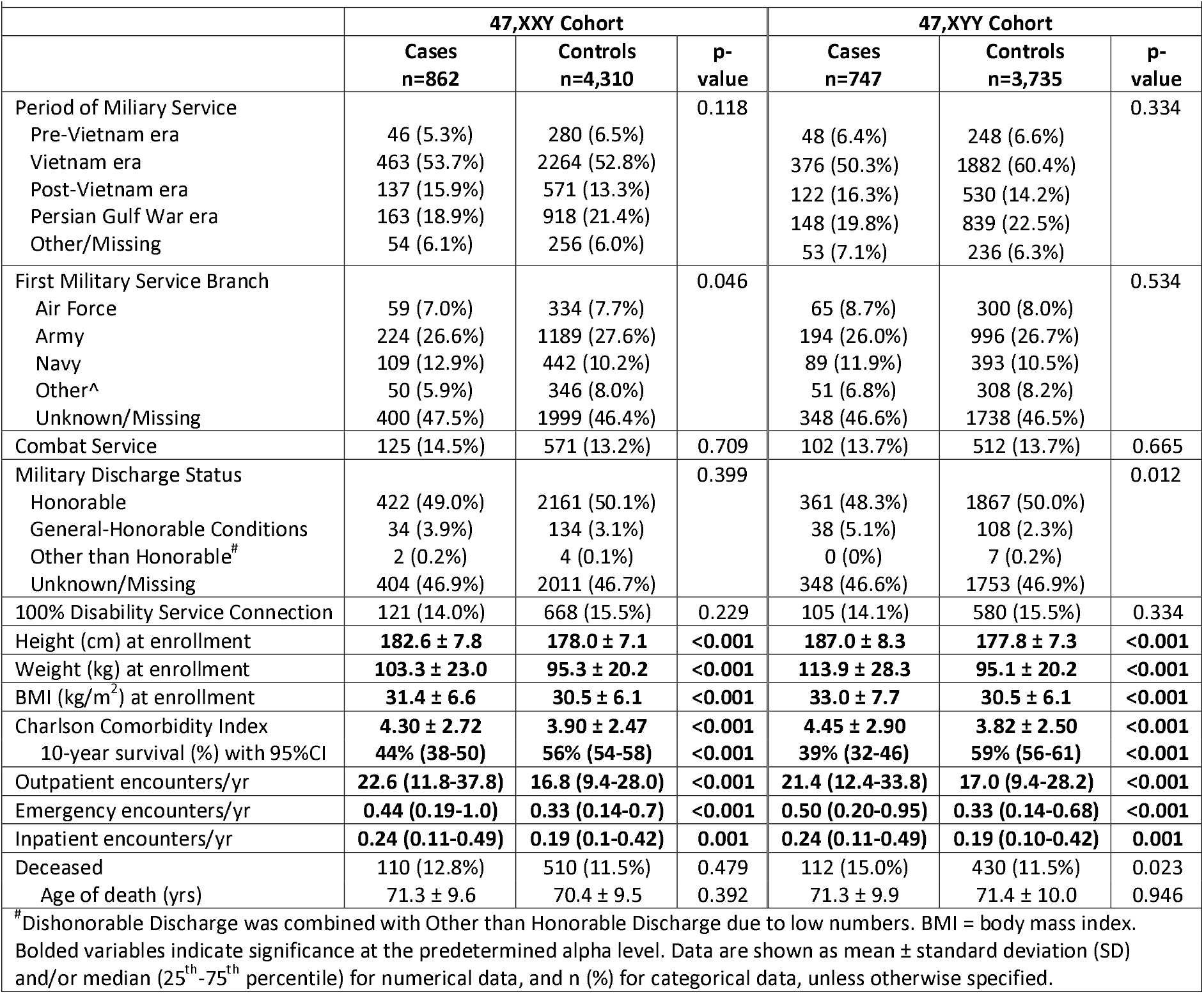
Miliary Service, Morbidity, and Mortality Assessed from VA Records in Men with 47,XXY or 47,XYY in the MVP and Matched Controls.

**Table 3.**
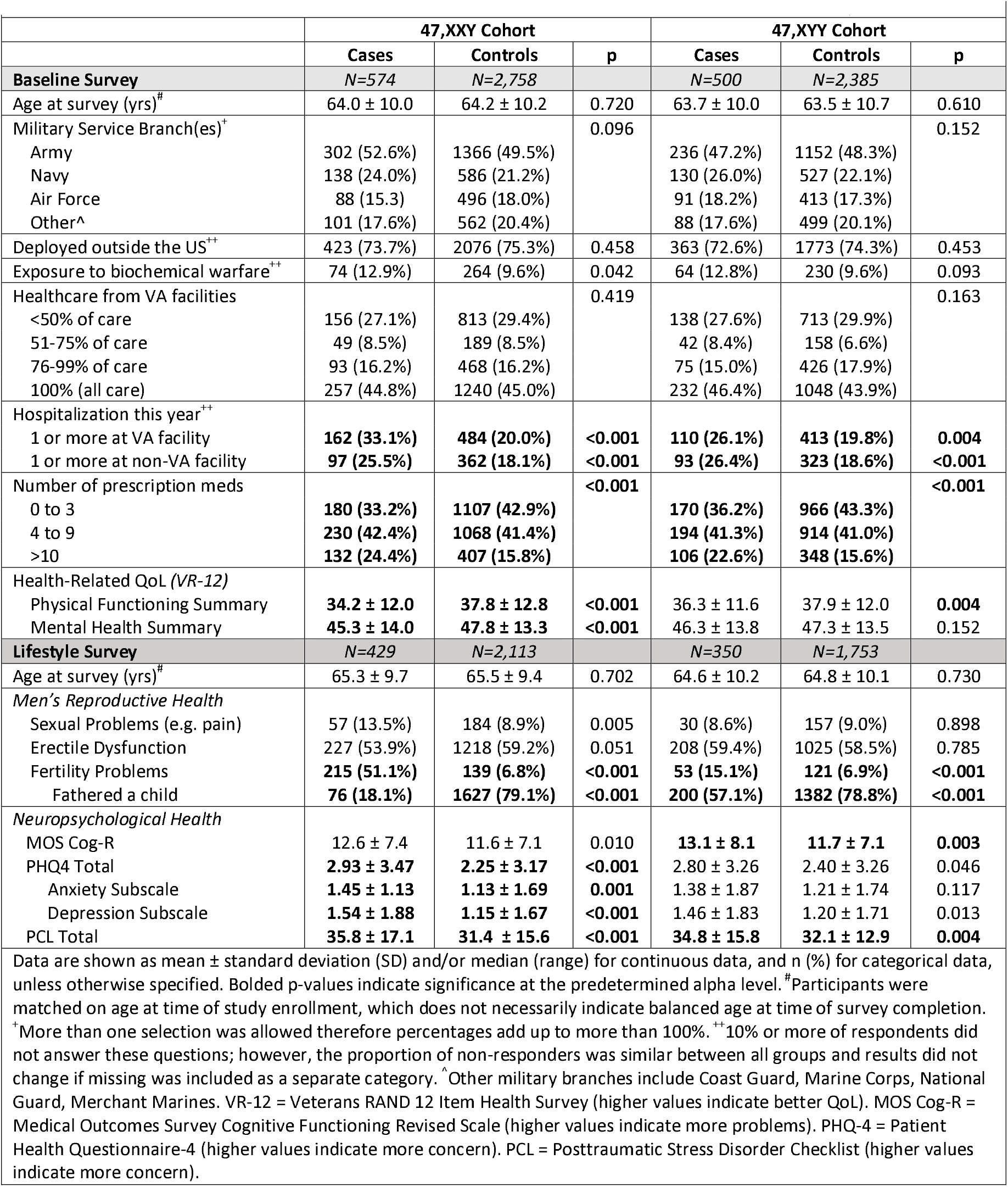
Self-Reported Military Service and Morbidity Metrics from the Baseline and Lifestyle MVP Surveys.

|  | 47,XXY Cohort |  |  | 47,XYY Cohort |  |  |
| --- | --- | --- | --- | --- | --- | --- |
|  | Cases | Controls | p | Cases | Controls | p |
| **Baseline Survey** | N=574 | N=2,758 |  | N=500 | N=2,385 |  |
| Age at survey (yrs)<sup>#</sup> | 64.0 ± 10.0 | 64.2 ± 10.2 | 0.720 | 63.7 ± 10.0 | 63.5 ± 10.7 | 0.610 |
| Military Service Branch(es)<sup>+</sup> |  |  | 0.096 |  |  | 0.152 |
| Army | 302 (52.6%) | 1366 (49.5%) |  | 236 (47.2%) | 1152 (48.3%) |  |
| Navy | 138 (24.0%) | 586 (21.2%) |  | 130 (26.0%) | 527 (22.1%) |  |
| Air Force | 88 (15.3) | 496 (18.0%) |  | 91 (18.2%) | 413 (17.3%) |  |
| Other<sup>^</sup> | 101 (17.6%) | 562 (20.4%) |  | 88 (17.6%) | 499 (20.1%) |  |
| Deployed outside the US<sup>**</sup> | 423 (73.7%) | 2076 (75.3%) | 0.458 | 363 (72.6%) | 1773 (74.3%) | 0.453 |
| Exposure to biochemical warfare<sup>**</sup> | 74 (12.9%) | 264 (9.6%) | 0.042 | 64 (12.8%) | 230 (9.6%) | 0.093 |
| Healthcare from VA facilities |  |  | 0.419 |  |  | 0.163 |
| <50% of care | 156 (27.1%) | 813 (29.4%) |  | 138 (27.6%) | 713 (29.9%) |  |
| 51-75% of care | 49 (8.5%) | 189 (8.5%) |  | 42 (8.4%) | 158 (6.6%) |  |
| 76-99% of care | 93 (16.2%) | 468 (16.2%) |  | 75 (15.0%) | 426 (17.9%) |  |
| 100% (all care) | 257 (44.8%) | 1240 (45.0%) |  | 232 (46.4%) | 1048 (43.9%) |  |
| Hospitalization this year<sup>**</sup> |  |  |  |  |  |  |
| 1 or more at VA facility | **162 (33.1%)** | **484 (20.0%)** | **<0.001** | **110 (26.1%)** | **413 (19.8%)** | **0.004** |
| 1 or more at non-VA facility | **97 (25.5%)** | **362 (18.1%)** | **<0.001** | **93 (26.4%)** | **323 (18.6%)** | **<0.001** |
| Number of prescription meds |  |  | **<0.001** |  |  | **<0.001** |
| 0 to 3 | **180 (33.2%)** | **1107 (42.9%)** |  | **170 (36.2%)** | **966 (43.3%)** |  |
| 4 to 9 | **230 (42.4%)** | **1068 (41.4%)** |  | **194 (41.3%)** | **914 (41.0%)** |  |
| >10 | **132 (24.4%)** | **407 (15.8%)** |  | **106 (22.6%)** | **348 (15.6%)** |  |
| Health-Related QoL (VR-12) |  |  |  |  |  |  |
| Physical Functioning Summary | **34.2 ± 12.0** | **37.8 ± 12.8** | **<0.001** | 36.3 ± 11.6 | 37.9 ± 12.0 | **0.004** |
| Mental Health Summary | **45.3 ± 14.0** | **47.8 ± 13.3** | **<0.001** | 46.3 ± 13.8 | 47.3 ± 13.5 | 0.152 |
| **Lifestyle Survey** | N=429 | N=2,113 |  | N=350 | N=1,753 |  |
| Age at survey (yrs)<sup>#</sup> | 65.3 ± 9.7 | 65.5 ± 9.4 | 0.702 | 64.6 ± 10.2 | 64.8 ± 10.1 | 0.730 |
| *Men's Reproductive Health* |  |  |  |  |  |  |
| Sexual Problems (e.g. pain) | 57 (13.5%) | 184 (8.9%) | 0.005 | 30 (8.6%) | 157 (9.0%) | 0.898 |
| Erectile Dysfunction | 227 (53.9%) | 1218 (59.2%) | 0.051 | 208 (59.4%) | 1025 (58.5%) | 0.785 |
| Fertility Problems | **215 (51.1%)** | **139 (6.8%)** | **<0.001** | **53 (15.1%)** | **121 (6.9%)** | **<0.001** |
| Fathered a child | **76 (18.1%)** | **1627 (79.1%)** | **<0.001** | **200 (57.1%)** | **1382 (78.8%)** | **<0.001** |
| *Neuropsychological Health* |  |  |  |  |  |  |
| MOS Cog-R | 12.6 ± 7.4 | 11.6 ± 7.1 | 0.010 | **13.1 ± 8.1** | **11.7 ± 7.1** | **0.003** |
| PHQ4 Total | **2.93 ± 3.47** | **2.25 ± 3.17** | **<0.001** | 2.80 ± 3.26 | 2.40 ± 3.26 | 0.046 |
| Anxiety Subscale | **1.45 ± 1.13** | **1.13 ± 1.69** | **0.001** | 1.38 ± 1.87 | 1.21 ± 1.74 | 0.117 |
| Depression Subscale | **1.54 ± 1.88** | **1.15 ± 1.67** | **<0.001** | 1.46 ± 1.83 | 1.20 ± 1.71 | 0.013 |
| PCL Total | **35.8 ± 17.1** | **31.4 ± 15.6** | **<0.001** | **34.8 ± 15.8** | **32.1 ± 12.9** | **0.004** |

Data are shown as mean ± standard deviation (SD) and/or median (range) for continuous data, and n (%) for categorical data, unless otherwise specified. Bolded p-values indicate significance at the predetermined alpha level. <sup>#</sup>Participants were matched on age at time of study enrollment, which does not necessarily indicate balanced age at time of survey completion. <sup>+</sup>More than one selection was allowed therefore percentages add up to more than 100%. <sup>\*\*</sup>10% or more of respondents did not answer these questions; however, the proportion of non-responders was similar between all groups and results did not change if missing was included as a separate category. <sup>^</sup>Other military branches include Coast Guard, Marine Corps, National Guard, Merchant Marines. VR-12 = Veterans RAND 12 Item Health Survey (higher values indicate better QoL). MOS Cog-R = Medical Outcomes Survey Cognitive Functioning Revised Scale (higher values indicate more problems). PHQ-4 = Patient Health Questionnaire-4 (higher values indicate more concern). PCL = Posttraumatic Stress Disorder Checklist (higher values indicate more concern).

Between enrollment and last follow up, 110 individuals with 47,XXY (12.8%) and 112 with 47,XYY (15%) died, compared to 11.5% of age-matched controls, yielding an SMR of 1.08 (95%CI 0.9-1.3) for 47,XXY and 1.30 (95%CI 1.1-1.6) for 47,XYY. Mean age of death for all groups was between 70-72 years with no statistical differences. Supplemental Table 1 details primary causes of death.

Men who were clinically diagnosed with Klinefelter syndrome (at an average age of 52) were younger with a lower CCI and more clinical encounters than those with 47,XXY on genotyping without a Klinefelter syndrome diagnosis in their EHR, (Supplemental Table 2). All other outcome measures from the EHR and from surveys were very similar between men with 47,XXY with and without a clinical diagnosis.

## DISCUSSION

In this large, ethnically diverse population-based study, we found a prevalence of 270 per 100,000 for X or Y chromosome aneuploidy among male veterans, which is similar or even higher than previous prevalence estimates in other populations.^1^ These men successfully completed military service many years prior to study enrollment with similar service metrics to their 46,XY peers, however, they experienced higher medical morbidity and lower HRQoL with aging. Evidence of a clinical diagnosis of 47,XXY was associated with lower medical comorbidity than those undiagnosed. An additional novel finding of this study was that the prevalence of SCA differed by genetic ancestry with half as many affected from African and Hispanic ancestries compared to those of European or East Asian ancestry. This initial overview of men with 47,XXY and 47,XYY within the MVP highlights the exciting potential of the MVP as a unique data source for investigating the effects of SCA on health outcome and quality of life during aging.

Prior epidemiologic studies, including birth cohorts and several population-based cohorts, have estimated that 47,XXY occurs in 103-174/100,000 males and 47,XYY in 69-98/100,000 males.^1,23^ Our estimates in the MVP population are within the expected range for 47,XXY (145 per 100,0000) and slightly higher than previously reported for 47,XYY (125 in 100,000). Not surprisingly, the minority of these individuals had a clinical diagnosis in the EHR, and the age of diagnosis in this cohort was much older than previously reported.^24^ If men with a SCA had been formally diagnosed prior to enlistment, certain Department of Defense codes regarding medical, learning, and psychiatric disorders could have prevented them from serving in the US military. Based on our study, however, the observed prevalence rates, along with comparable service metrics, provide evidence that men with SCAs are as capable of successful military service as their 46,XY peers.

Our study represents one of the only estimations of the prevalence of 47,XXY and 47,XYY among men in a US population, and the only one to our knowledge inclusive of diverse ancestral groups. We did not expect to see a difference in prevalence based on ancestry, as the risk of chromosomal aneuploidy is assumed to be independent of genetic or social constructs because it is caused by random errors in meiosis.^25^ Parental age could be a confounder, as nondisjunction errors increase with age and parental age is higher in European ancestry compared to Hispanic or African ancestry.^26^ However, the role of paternal age in aneuploidy risk is less clear than that of maternal age,^27^ and while there was a significant difference in parental age between men with 47,XXY and controls, that difference was not present between men with 47,XYY and controls. Alternatively, the cross-ancestry differences in prevalence could be secondary to sociodemographic factors. Enlistment in the US military requires conformation to a system of expectations that may be less attainable for minoritized individuals who also have physical, socioemotional, and learning differences that can be associated with SCAs^6^ – essentially a double hit. Undoubtedly, further research is needed to understand the basis of the ancestorial differences in the prevalence of SCAs, including prevalence estimates in non-Western countries and other diverse populations.

Our results corroborate findings from multiple other studies that a supernumerary sex chromosome is associated with excess morbidity and higher healthcare utilization.^2,4,6^ It is promising that a clinical diagnosis of 47,XXY, even if quite delayed, was associated with a lower CCI, possibly due to closer medical follow-up and more preventive medicine interventions in the VA Healthcare System. Future studies should examine whether the relationship between clinical diagnosis, increased outpatient encounters, and reduced morbidity is causal. Additional investigation into the specific medical conditions contributing to the increased morbidity index is underway.

We also observed poorer self-reported health and wellbeing metrics among men with SCA. Lower QoL outcomes are widely reported in SCAs, however, to our knowledge this is the only study that has evaluated HRQoL in individuals who are presumably unaware of their genetic condition. Biased stereotypes for 47,XXY and 47,XYY can continue to influence expectations and reporting for subjective measures, both by participants and researchers, highlighting the valuable contribution of our findings to the existing literature. QoL outcomes did not differ between those with and without a clinical diagnosis in our study, suggesting a true association between SCA and QoL rather than clinical ascertainment as a confounder. In one study, health status was the most significant predictor of QoL in men with 47,XXY,^28^ suggesting QoL may be increased if chronic health were improved. In addition, many conditions contributing to morbidity in SCAs are preventable or at least treatable.^6,29^ We hypothesize that a more timely diagnosis could optimize medical care, improve health, and ultimately lead to better QoL with aging.

In contrast to studies from Europe, we did not find greater mortality in men with SCAs.^3,5,30^ This may reflect a limitation of the MVP dataset as we cannot capture deaths occurring prior to enrollment (mean age of 61 years), and the follow up duration is relatively short for mortality disparities to emerge. Given the estimated 10-year survival was significantly lower in SCA groups, longitudinal follow up will be important to clarify the impact of SCAs on mortality. In addition, while the life expectancy of men with SCA in the MVP was similar to prior reports (∼71 years), our controls did not live as long as men in the European cohorts, possibly reflecting poorer health among US Veterans.^31^

This is the first US, ancestrally diverse investigation of SCAs, with rigorous cohort identification through genotyping that overcomes the clinical diagnostic bias inherent in most SCA studies to date. Despite these strengths, we caution generalizing these findings to other populations. Young men with a known SCA diagnosis may have been dissuaded from enrolling in the military, and VHA clinicians may be less likely to suspect and test for a genetic condition in men who have served in the military (reverse diagnostic suspicion bias), affecting the prevalence and diagnostic estimates within. We relied on the VHA EHR to determine if an individual had a clinical diagnosis of 47,XXY or 47,XYY, which may underestimate the true diagnostic rate if a known diagnosis was withheld from the EHR for any reason, including patient non-disclosure. Furthermore, while the MVP cohort has provided a unique opportunity to study an aging population with men with SCAs, earlier outcomes may not be captured. Finally, although the rate of survey response was congruent with the MVP cohort as a whole (67%), it is possible that respondents may not be representative of the non-responders with 47,XXY/47,XYY. Recognizing these limitations, this robust study provides important, novel data for the SCA community and introduces the potential of this valuable resource.

## CONCLUSIONS

In the largest and most ethnically diverse population-based study of 47,XXY and 47,XYY to date, we observed 270 out of 100,000 men in the MVP study has an extra X or Y chromosome, with an unexpected finding of a higher prevalence among European and East Asian ancestry groups. Our analysis establishes that men with SCA successfully complete military duty with similar service performance metrics to their 46,XY peers. Thus, we urge that neither these conditions nor their treatments be exclusionary to military service. Although several multiple health and QoL are negatively impacted by the presence of an additional X or Y chromosome, these results provide evidence for the first time that a clinical diagnosis may reduce comorbidity burden. These data add to the existing literature on male SCA with a favorable perspective on their contribution to society, and highlight the need to study preventive medicine and other interventions that may ameliorate the observed negative health sequelae observed during aging.

## Data Availability

MVP genomic and phenotypic data are currently available to VA investigators and approved partners, with plans for expanding to non-VA investigators in the future. Contact for questions related to data access.

https://www.mvp.va.gov/pwa/discover-mvp-data

## ACKNOWLEDGEMENTS

This research used data from the Million Veteran Program (MVP) [Office of Research and Development, Veterans Health Administration] and was supported by MVP022 award CX001727 (PI: Hauger) and MVP000 award (PI: Pyarajan). RLH was additionally funded by the VISN-22 VA Center of Excellence for Stress and Mental Health (CESAMH) and National Institute of Aging R01 grants AG050595, AG05064, AG065385. SMD received salary support during this work from NICHD K23HD092588. MSP was supported on awards 1F30CA247168-01 and T32CA067754. VCM received salary support during this work from IK2 CX001952 from the VA Clinical Science Research & Development Service. This publication does not represent the views of the Department of Veterans Affairs, the National Institutes of Health, or the United States Government.

## REFERENCES

1. Berglund A, Stochholm K, Gravholt CH. The epidemiology of sex chromosome abnormalities. American journal of medical genetics Part C, Seminars in medical genetics. Jun 2020;184(2):202–215. doi:10.1002/ajmg.c.31805

2. Berglund A, Stochholm K, Gravholt CH. Morbidity in 47,XYY syndrome: a nationwide epidemiological study of hospital diagnoses and medication use. Genetics in medicine : official journal of the American College of Medical Genetics. Sep 2020;22(9):1542–1551. doi:10.1038/s41436-020-0837-y

3. Stochholm K, Juul S, Gravholt CH. Diagnosis and mortality in 47,XYY persons: a registry study. Orphanet journal of rare diseases. 2010;5:15. doi:10.1186/1750-1172-5-15

4. Bojesen A, Gravholt CH. Morbidity and mortality in Klinefelter syndrome (47,XXY). Acta paediatrica. Jun 2011;100(6):807–13. doi:10.1111/j.1651-2227.2011.02274.x

5. Higgins CD, Swerdlow AJ, Schoemaker MJ, Wright AF, Jacobs PA, Group UKCC. Mortality and cancer incidence in males with Y polysomy in Britain: a cohort study. Human genetics. Jul 2007;121(6):691–6. doi:10.1007/s00439-007-0365-8

6. Ridder LO, Berglund A, Stochholm K, Chang S, Gravholt CH. Morbidity, mortality, and socioeconomics in Klinefelter syndrome and 47,XYY syndrome: a comparative review. Endocr Connect. May 1 2023;12(5)doi:10.1530/EC-23-0024

7. Mehmet B, Gillard S, Jayasena CN, Llahana S. Association between domains of quality of life and patients with Klinefelter syndrome: a systematic review. European journal of endocrinology / European Federation of Endocrine Societies. Aug 1 2022;187(2):S21–S34. doi:10.1530/EJE-21-1239

8. Zhao Y, Gardner EJ, Tuke MA, et al. Detection and characterization of male sex chromosome abnormalities in the UK Biobank study. Genetics in medicine : official journal of the American College of Medical Genetics. Sep 2022;24(9):1909–1919. doi:10.1016/j.gim.2022.05.011

9. Gaziano JM, Concato J, Brophy M, et al. Million Veteran Program: A mega-biobank to study genetic influences on health and disease. J Clin Epidemiol. Feb 2016;70:214–23. doi:10.1016/j.jclinepi.2015.09.016

10. Harrington KM, Nguyen XT, Song RJ, et al. Gender Differences in Demographic and Health Characteristics of the Million Veteran Program Cohort. Womens Health Issues. Jun 25 2019;29 Suppl 1(Suppl 1):S56–S66. doi:10.1016/j.whi.2019.04.012

11. Nguyen XT, Quaden RM, Song RJ, et al. Baseline Characterization and Annual Trends of Body Mass Index for a Mega-Biobank Cohort of US Veterans 2011-2017. J Health Res Rev Dev Ctries. 2018;5(2):98–107.

12. Hunter-Zinck H, Shi Y, Li M, et al. Genotyping Array Design and Data Quality Control in the Million Veteran Program. American journal of human genetics. Apr 2 2020;106(4):535–548. doi:10.1016/j.ajhg.2020.03.004

13. Fang H, Hui Q, Lynch J, et al. Harmonizing Genetic Ancestry and Self-identified Race/Ethnicity in Genome-wide Association Studies. American journal of human genetics. Oct 3 2019;105(4):763–772. doi:10.1016/j.ajhg.2019.08.012

14. Gaspar HA, Breen G. Probabilistic ancestry maps: a method to assess and visualize population substructures in genetics. BMC Bioinformatics. Mar 7 2019;20(1):116. doi:10.1186/s12859-019-2680-1

15. Charlson ME, Pompei P, Ales KL, MacKenzie CR. A new method of classifying prognostic comorbidity in longitudinal studies: development and validation. J Chronic Dis. 1987;40(5):373–83. doi:10.1016/0021-9681(87)90171-8

16. Kazis LE, Miller DR, Skinner KM, et al. Applications of methodologies of the Veterans Health Study in the VA healthcare system: conclusions and summary. J Ambul Care Manage. Apr-Jun 2006;29(2):182–8. doi:10.1097/00004479-200604000-00011

17. Clark AL, McGill MB, Ozturk ED, et al. Self-reported physical functioning, cardiometabolic health conditions, and health care utilization patterns in Million Veteran Program enrollees with Traumatic Brain Injury Screening and Evaluation Program data. Mil Med Res. Jan 3 2023;10(1):2. doi:10.1186/s40779-022-00435-7

18. Merritt VC, Crocker LD, Sakamoto MS, Chanfreau-Coffinier C, Delano-Wood L, Million Veteran P. Psychiatric symptoms influence social support in VA Million Veteran Program enrollees screening positive for traumatic brain injury. Soc Sci Med. Nov 2022;312:115372. doi:10.1016/j.socscimed.2022.115372

19. Fink SJ, Davey DK, Sakamoto MS, et al. Subjective cognitive and psychiatric well-being in U.S. Military Veterans screened for deployment-related traumatic brain injury: A Million Veteran Program Study. Journal of psychiatric research. Jul 2022;151:144–149. doi:10.1016/j.jpsychires.2022.04.019

20. Kroenke K, Spitzer RL, Williams JB, Lowe B. An ultra-brief screening scale for anxiety and depression: the PHQ-4. Psychosomatics. Nov-Dec 2009;50(6):613–21. doi:10.1176/appi.psy.50.6.613

21. Blevins CA, Weathers FW, Davis MT, Witte TK, Domino JL. The Posttraumatic Stress Disorder Checklist for DSM-5 (PCL-5): Development and Initial Psychometric Evaluation. J Trauma Stress. Dec 2015;28(6):489–98. doi:10.1002/jts.22059

22. Instructions for classifying the underlying cause of death.

23. Coffee B, Keith K, Albizua I, et al. Incidence of fragile X syndrome by newborn screening for methylated FMR1 DNA. American journal of human genetics. Oct 2009;85(4):503–14. doi:10.1016/j.ajhg.2009.09.007

24. Berglund A, Viuff MH, Skakkebaek A, Chang S, Stochholm K, Gravholt CH. Changes in the cohort composition of turner syndrome and severe non-diagnosis of Klinefelter, 47,XXX and 47,XYY syndrome: a nationwide cohort study. Orphanet journal of rare diseases. Jan 14 2019;14(1):16. doi:10.1186/s13023-018-0976-2

25. Griffin DK. The incidence, origin, and etiology of aneuploidy. Int Rev Cytol. 1996;167:263–96. doi:10.1016/s0074-7696(08)61349-2

26. Subramanian VV, Bickel SE. Aging predisposes oocytes to meiotic nondisjunction when the cohesin subunit SMC1 is reduced. PLoS Genet. Nov 2008;4(11):e1000263. doi:10.1371/journal.pgen.1000263

27. Fonseka KG, Griffin DK. Is there a paternal age effect for aneuploidy? Cytogenetic and genome research. 2011;133(2-4):280–91. doi:10.1159/000322816

28. Rapp M, Mueller-Godeffroy E, Lee P, et al. Multicentre cross-sectional clinical evaluation study about quality of life in adults with disorders/differences of sex development (DSD) compared to country specific reference populations (dsd-LIFE). Health Qual Life Outcomes. Apr 3 2018;16(1):54. doi:10.1186/s12955-018-0881-3

29. Chang S, Skakkebaek A, Davis SM, Gravholt CH. Morbidity in Klinefelter syndrome and the effect of testosterone treatment. American journal of medical genetics Part C, Seminars in medical genetics. Jun 2020;184(2):344–355. doi:10.1002/ajmg.c.31798

30. Swerdlow AJ, Higgins CD, Schoemaker MJ, Wright AF, Jacobs PA, United Kingdom Clinical Cytogenetics G. Mortality in patients with Klinefelter syndrome in Britain: a cohort study. The Journal of clinical endocrinology and metabolism. Dec 2005;90(12):6516–22. doi:10.1210/jc.2005-1077

31. Schult TM, Schmunk SK, Marzolf JR, Mohr DC. The Health Status of Veteran Employees Compared to Civilian Employees in Veterans Health Administration. Mil Med. Jul 1 2019;184(7-8):e218–e224. doi:10.1093/milmed/usy410

