## Supplemental Tables for "Prevalence, Morbidity, and Mortality of 1,609 Men with Sex Chromosome Aneuploidy: Results from the Diverse Million Veteran Program Cohort"

Supplemental Material

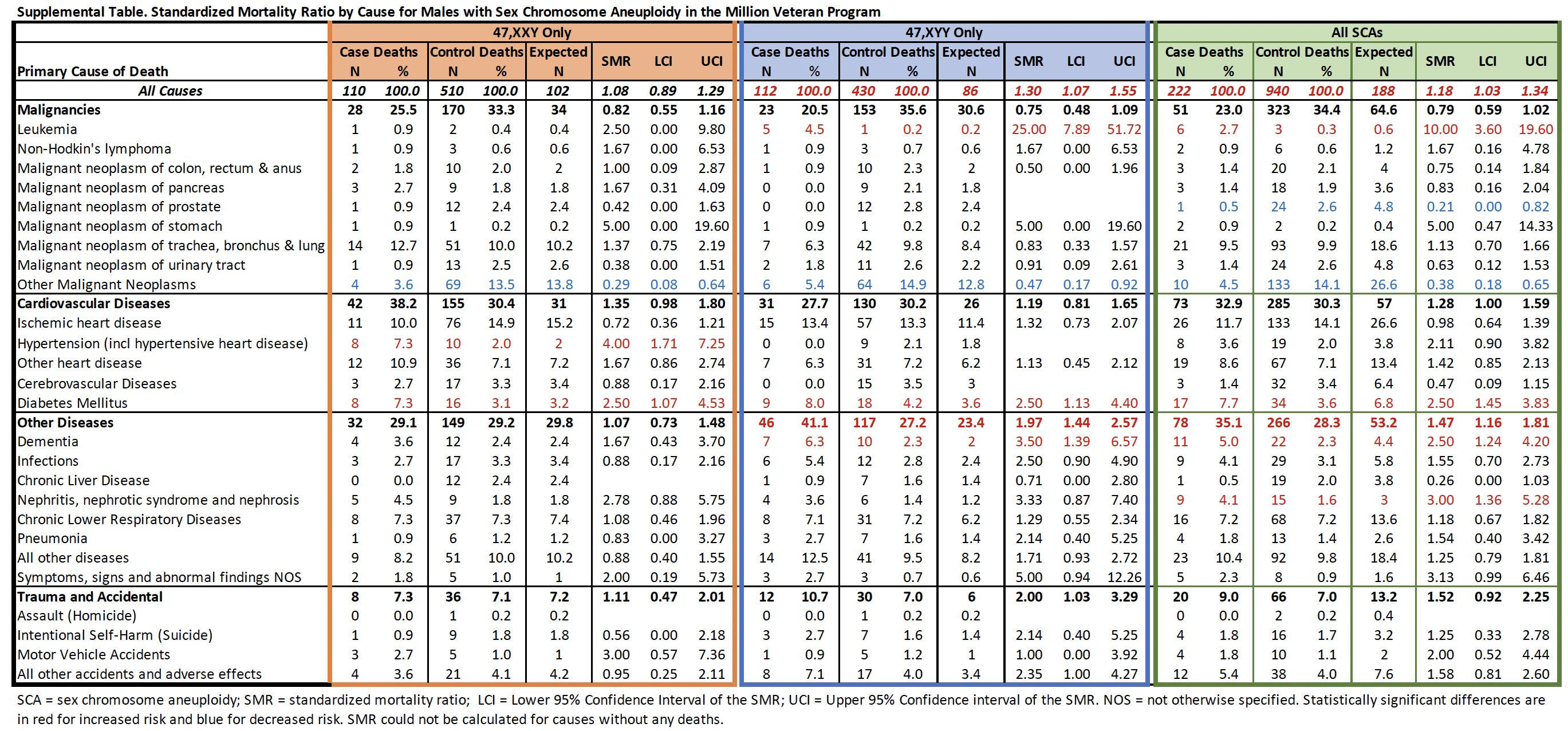

| **Supplemental Table 2. 47,XXY Stratified by Clinical Diagnosis** | | | | | | | |
| --- | --- | --- | --- | --- | --- | --- | --- |
|  | ***Clinically Diagnosed Cohort*** | | | ***Undiagnosed Cohort*** | | | **47,XXY diagnosed vs undiagnosed p-value** |
|  | **Diagnosed Cases Total N=226 BL survey n=137 Life survey n=88** | **Controls Total N=1130 BL survey n=687 Life survey n=475** | **cases vs controls p-value** | **Undiagnosed Cases Total N=640 BL survey n=437 Life survey n=333** | **Controls Total N=3200 BL survey n=2092 Life survey n=1560** | **cases vs controls p-value** |  |
| **Demographics and Military Service** |  |  |  |  |  |  |  |
| Age at enrollment (yrs) | 56.9 ± 13.4 | 56.9 ± 13.4 | 1 | 62.7 ± 11.2 | 62.7 ± 11.2 | 1 | **<0.001** |
| European Genetic Ancestry | 190 (84.1%) | 950 (84.1%) | 1 | 539 (84.2%) | 2695 (84.2%) | 1 | 0.958 |
| Service Era: Vietnam | 91 (40.2%) | 456 (40.4%) | 0.980 | 373 (58.3%) | 1847 (57.7%) | 0.793 | **<0.001** |
| Military Branch: Army* | 64 (50.0%) | 312 (50.4%) | 0.722 | 180 (53.9%) | 869 (51.7%) | 0.036 | 0.959 |
| Combat Service: Yes* | 45 (21.3%) | 137 (14.5%) | **<0.001** | 80 (12.5%) | 410 (12.8%) | 0.124 | **0.005** |
| Deployed outside the US | 96 (70.1%) | 493 (71.8%) | 0.680 | 329 (74.8%) | 1585 (75.8%) | 0.669 | 0.317 |
| Exposure to biochemical warfare | 24 (17.5%) | 58 (8.4%) | **0.009** | 50 (11.4%) | 200 (9.6%) | 0.420 | 0.200 |
| Honorable Discharge* | 120 (95.2%) | 578 (93.8%) | 0.776 | 303 (91.0%) | 1566 (93.7%) | 0.202 | 0.184 |
| **Healthcare Utilization** |  |  |  |  |  |  |  |
| Care within VA system: 100% | 68 (49.6%) | 289 (42.1%) | **0.004** | 189 (43.0%) | 912 (43.6%) | 0.485 | **0.009** |
| Outpatient encounters/yr | 26.6 (14.9-43.2) | 16.7 (9.3-28.0) | **<0.001** | 22.2 (11.3-36.0) | 17.2 (9.6-28.3) | **<0.001** | **0.001** |
| Emergency encounters/yr | 0.46 (0.21-1.0) | 0.37 (0.15-0.77) | **0.005** | 0.26 (0.05-0.76) | 0.33 (0.14-0.67) | **<0.001** | **<0.001** |
| Inpatient encounters/yr | 0.11 (0.0-0.33) | 0.05 (0.0-0.24) | **<0.001** | 0.13 (0.0-0.33) | 0.06 (0.0-0.24) | **<0.001** | 0.455 |
| Hospitalized in past year (VA) | 40 (29.2%) | 113 (16.5%) | **<0.001** | 122 (27.9%) | 359 (17.2%) | **<0.001** | 0.642 |
| **Medical Characteristics & Morbidity** |  |  |  |  |  |  |  |
| Height (cm) at enrollment | 182.7 ± 8.4 | 178.4 ± 7.0 | **<0.001** | 182.5 ± 7.7 | 178.0 ± 7.2 | **<0.001** | 0.822 |
| Weight (kg) at enrollment | 103.3 ± 21.9 | 96.1 ± 20.1 | **<0.001** | 103.1 ± 23.4 | 94.9 ± 20.1 | **<0.001** | 0.902 |
| BMI (kg/m2) at enrollment | 31.4 ± 6.2 | 30.8 ± 6.2 | 0.153 | 31.4 ± 6.8 | 30.4 ± 6.1 | **<0.001** | 0.954 |
| Charlson Comorbidity Index | 3.65 ± 2.73 | 3.39 ± 2.55 | 0.198 | 4.53 ± 2.68 | 4.13 ± 2.47 | **<0.001** | **<0.001** |
| >10 Prescription Medications | 35 (25.5%) | 95 (13.8%) | **<0.001** | 97 (22.0%) | 311 (14.9%) | **<0.001** | 0.052 |
| Sexual Health Problems | 18 (20.5%) | 40 (8.4%) | **0.002** | 39 (11.6%) | 152 (9.7%) | 0.272 | **0.036** |
| Fertility Problems | 52 (59.1%) | 31 (6.5%) | **<0.001** | 164 (48.8%) | 110 (7.1%) | **<0.001** | 0.094 |
| **Psychosocial Morbidity** |  |  |  |  |  |  |  |
| VR-12 Physical Functioning | **34.8 ± 12.4** | **38.4 ± 13.3** | **0.005** | **34.0 ± 11.9** | **37.6 ± 12.8** | **<0.001** | 0.519 |
| VR-12 Mental Health | **42.8 ± 15.0** | **46.5 ± 14.0** | **0.009** | **46.0 ± 13.6** | **48.2 ± 13.2** | **0.003** | **0.023** |
| MOS Cog-R | 12.6 ± 6.0 | 12.2 ± 7.6 | 0.608 | 12.6 ± 7.7 | 11.5 ± 7.1 | **0.020** | 0.985 |
| PHQ4 Total | 3.14 ± 3.4 | 2.45 ± 3.29 | 0.084 | 2.87 ± 3.5 | 2.23 ± 3.2 | **0.002** | 0.487 |
| PHQ4 Anxiety Subscale | 1.54 ± 1.87 | 1.19 ± 1.7 | 0.106 | 1.42 ± 1.86 | 1.12 ± 1.7 | **0.005** | 0.598 |
| PHQ4 Depression Subscale | 1.60 ± 1.78 | 1.29 ± 1.78 | 0.128 | 1.51 ± 1.91 | 1.14 ± 1.68 | **<0.001** | 0.657 |
| PCL Total | 36.5 ± 17.0 | 32.6 ± 16.3 | 0.055 | 35.6 ± 17.1 | 31.5 ± 15.7 | **<0.001** | 0.649 |
| **Mortality** |  |  |  |  |  |  |  |
| Deceased | 20 (8.8%) | 127 (11.2%) | 0.292 | 90 (14.1%) | 410 (12.8%) | 0.493 | 0.092 |
| Colored text indicates the data source and subsequently the sample size available for that variable: black is the electronic health record database, blue is the Baseline Survey, and green is the Lifestyle Survey. Data are shown as mean ± standard deviation (SD) and/or median (range) for continuous data, and n (%) for categorical data, unless otherwise specified. Bolded p-values indicate statistical significance at an alpha of 0.05. *Percentages are calculated from the total with non-missing data; chi-squared analyses included missing as a category. VA = Veterans Affairs; VR-12 = Veterans RAND 12 Item Health Survey (higher values indicate better QoL). MOS Cog-R = Medical Outcomes Survey Cognitive Functioning Revised Scale (higher values indicate more problems). PHQ-4 = Patient Health Questionnaire-4 (higher values indicate more concern). PCL = Posttraumatic Stress Disorder Checklist (higher values indicate more concern). | | | | | | | |
